# Intermittent radiotherapy as alternative treatment for recurrent high grade glioma: A modelling study based on longitudinal tumor measurements

**DOI:** 10.1101/2021.01.09.21249317

**Authors:** Sarah C. Brüningk, Jeffrey Peacock, Christopher J. Whelan, Hsiang-Hsuan M. Yu, Solmaz Sahebjam, Heiko Enderling

## Abstract

Recurrent high grade glioma patients face a poor prognosis for which no curative treatment option currently exists. In contrast to prescribing high dose hypofractionated stereotactic radiotherapy (HFSRT, ≥ 6 *Gy*x5 in daily fractions) with debulking intent, we suggest a personalized treatment strategy to improve tumor control by delivering intermittent high dose treatment (iRT, ≥ 6 *Gy*x1 every six weeks). We performed a simulation analysis to compare HFSRT, iRT and iRT plus boost (≥ 6 *Gy*x3 in daily fractions at time of progression) based on a mathematical model of tumour growth, radiation response and patient-specific evolution of resistance to additional treatments (pembrolizumab and bevacizumab). Model parameters were fitted from tumour growth curves of 16 patients enrolled in the phase 1 NCT02313272 trial that combined HFSRT with bevacizumab and pembrolizumab. Then, iRT +/-boost treatments were simulated and compared to HFSRT based on time to tumor regrowth. The modelling results demonstrated that iRT+boost(-boost) treatment was equal or superior to HFSRT in 15(11) out of 16 cases and that patients that remained responsive to pembrolizumab and bevacizumab would benefit most from iRT. Time to progression could be prolonged through the application of additional, intermittently delivered fractions. iRT hence provides a promising treatment option for recurrent high grade glioma patients.

## 1 Introduction

Patients with recurrent high-grade glioma (HGG), such as glioblastoma, face a dismal prognosis with median overall survival rates of less than one year^1,2^. This is likely related to the biological nature of these types of tumours which are characterised as fast growing, infiltrating, and frequently multifocal disease^3^. The diffuse nature of these tumors implies that any localized treatment, such as surgery or radiotherapy, inevitably fails to treat all (microscopic) disease and recurrences may hence occur either at the primary, or a distal location within the brain. According to NCCN guidlines^4^ there is no well defined standard of care for these patients and treatment options are limited. Hence, treatment strategy is often suggested on an individualised basis. These include re-resection of the tumour, systemic therapy such as bevacizumab, Lomustine, or Temozolomide, and palliative re-irradiation. Notably, re-irradiation in the recurrent HGG setting may be considered as a Category 2B option. Recently, alternative approaches incorporating immunotherapy^5^, have been tested for recurrent HGG in several clinical trials (see Laub *et al*. for an extensive review^6,7^) but the efficacy of this treatment could not be demonstrated. Inevitably, HGG tumors develop resistance to these systemic therapies.

In our recent phase 1 clinical trial (NCT02313272, 05/12/2014) recurrent HGG patients were treated with a combination of hypofractionated stereotactic radiotherapy (HFSRT; ≥ 6 *Gy* x 5 fractions), bevacizumab (antibody against vascular endothelial growth factor (VEGF)) and pembrolizumab (anti PD1 antibody)^8^. This study demonstrated safety in terms of adversarial side effects for this particular protocol. Although efficacy was not the primary endpoint, the response results were promising; yet median time to progression remained below one year^8^. In this trial, HFSRT was given in doses with maximum log cell kill intent over one week (consistent with current practice and trials for recurrent HGG re-irradiation studies^9^). While this dose fractionation strategy can be effective in eradicating cancer cells, it may select for radiation-resistant subclones by preferential killing of radio-sensitve subclones through an ecological-evolutionary process called competitive release^10–12^. The current protocol of maximum tolerable dose did not account for such evolutionary dynamics, and every patient inevitably developed resistance. Furthermore, HFSRT given upfront prevented the possibility to re-irradiate any additional (local or distant) recurrences, and provided only a single immune stimulus for anti PD1 treatment.

Here, we investigate through mathematical modelling an alternative, intermittent approach for radiotherapy treatment schedules. This approach is motivated by the assumption that for this group of patients tumor management will prolong time to progression compared to (failed) tumour eradication. The rational of evolutionary principles-guided intermittent treatment approach is to maintain a treatment-sensitive population that competes for resources with resistant cells and thus slows the expansion of a resistant clone, thereby prolonging time to progression^13,14^. Moreover, when delivering radiotherapy using an intermittent schedule, treatment dosing and irradiation volume can be adapted based on observed responses.

In this manuscript we describe a mathematical model, using only two patient specific parameters, that is suitable to fit clinically observed longitudinal, volumetric tumor growth in patients enrolled in the NCT02313272 trial. We use this model to simulate alternative, intermittent treatment schedules to determine whether these provide superior and personalisable alternatives to the current HFSRT for protocol for recurrent HGG.

## 2 Materials and Methods

### 2.1 Patient cohort

Patients with recurrent HGG included in this modelling study (n=16) were treated at the Moffitt Cancer Center, FL between August 2015 and March 2018 as part of a phase I clinical trial (NCT02313272, 05/12/2014)^8^. All patients provided written consent and the treatment protocol was approved by the institutional review board (IRB study #: Pro00014674 and # 00000971). Following optional surgical resection, all patients received HFSRT (≥6 *Gy*x5 delivered as five consecutive, daily fractions).

Here, treatment was prescribed as 30-35 Gy to the planning target volume (PTV) with a simultaneously integrated boost to the gross tumour volume (GTV) of *D*_95%_ = 30 − 40 Gy. All treatment plans were calculated in iPlan (Version 1.1 Brainlab, Munich, Germany) and were delivered as intensity modulated radiotherapy treatments using volumetric modulated arc therapy with image guidance. Planned doses were summarized in terms of generalised equivalent uniform dose (gEUD)^15,16^ and near minimum dose *D*_98%_, delivered to the PTV. gEUD ranged between 31.3 and 37.0 Gy, whereas the corresponding near minimum dose *D*_98%_ was between 28.5 and 35.9 Gy (see Table S1 for specific total dose per patient). gEUD calculations were performed in Matlab (version 2020a) using an exponent (Lyman parameter) of −10 as suggested previously^17^. The gEUD accounts for dose inhomogeneity, whereas the PTV captures geometric delivery uncertainties across the gross tumour volume, providing the basis for volumetric response evaluation.

In addition to HFSRT, all patients received the VEGF inhibitor bevacizumab (10 mg/kg, intravenously delivered every two weeks) and the anti PD1 antibody pembrolizumab (100 mg or 200 mg, intravenously applied every three weeks). Both, bevacizumab and pembrolizumab were given until time of progression (scored by a 20% increase in tumor volume above the nadir as per RANO criteria^18^) or toxicity. Tumour volume was assessed pre-treatment and approximately every six weeks (median 42 days, standard deviation 38 days) using T1-weighted, contrast enhanced magnetic resonance imaging (3T MRI, 1.5mm slice thickness). The region of hyperintensity on post-contrast T1-weighted MR images was contoured by a neuro-radiation oncologist as the GTV. Where required, additional MRI sequences such as T2-weighted and/or FLAIR imaging were used to accurately assess this GTV, especially when there was significant tumor associated edema. A 5 mm expansion was made from the GTV to create the PTV.

A subset of 16 trial patients (both bevacizumab naive and pretreated) with tumour measurements beyond the time of progression (i.e. tumour regrowth) was used in this study. Patients excluded from this analysis either left the trial due to reasons other than tumor progression, or their tumor regrowth was not quantified. For the selected group, four to ten (median six) post treatment data points were acquired. Figure 1 outlines the trial design (Figure 1A) and the patient subset included in this analysis (Figure 1B), and the patient characteristics (Figure 1C). Patients are shown with arbitrary identifies.

**Figure 1.**
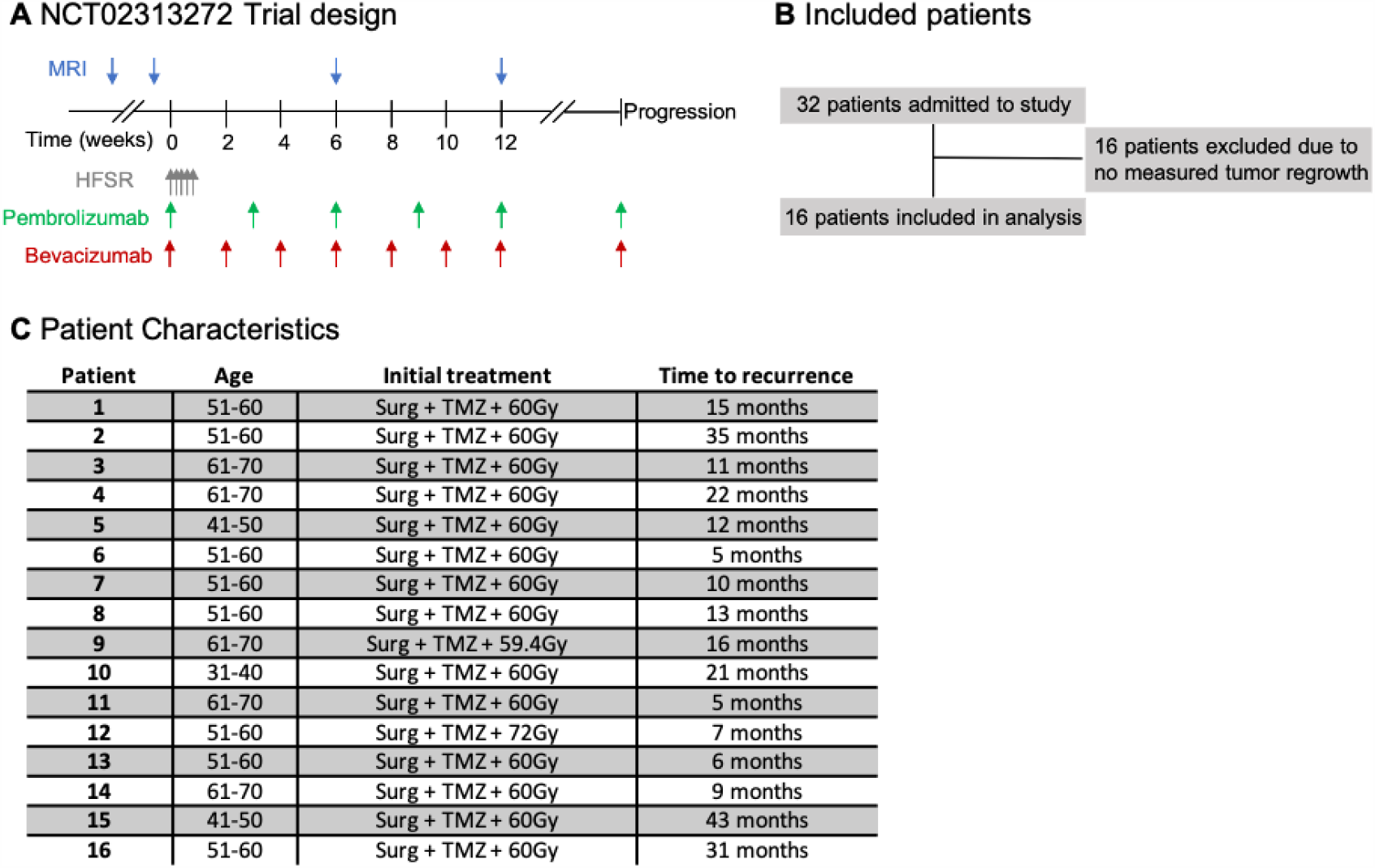
Overview of the used data. **A**: Schematic of the NCT02313272 protocol indicating the imaging and treatment time point for this triple combination therapy trial. **B**: Out of the 32 trial participants only those 16 with monitored tumour progression were included in this analysis. **C**: Characteristics of the 16 included patients. Abbreviations: HFSRT: Hypofractionated Stereotactic Radiosurgery, Surg: surgery, TMZ: temozolomide

### 2.2 Mathematical model

The aim of this study was to provide a simple mathematical framework to (i) fit the observed tumour growth response data to HFSRT given in five daily fractions, and, based on this description, to (ii) simulate intermittent radiation treatment (iRT) schedules. The presented model captures only the key mechanisms of treatment response to limit the mathematical complexity of the model to be able to obtain high confidence fit parameters estimates. We extended a mathematical tumor-growth inhibition model reported by Glazar *et al*. to account for the contribution of HFSRT to tumour volume reduction^19^. Tumor volume growth was described as exponential growth at rate *λ* [day^−1^], hence neglecting potential plateauing effects due to limited carrying capacity of a tumour^20^ within this time frame. Upon treatment initiation, the effect and onset of resistance to bevacizumab and pembrolizumab treatment was modelled as previously described^19^ by exponential tumor volume reduction with rate *γ* [day^−1^]:

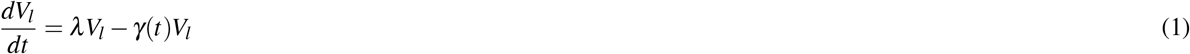

Here, *V*_*l*_(*t*) is the viable tumour volume at time *t*, and *γ*(*t*) denotes the volume decay due to Bevacizumab and Pembrolizumab treatment. As treatment resistance builds up, this decay rate exponentially decreases at a characteristic rate *ε*:

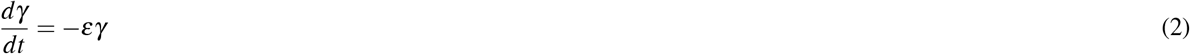

In summary this leads to the following analytic solution to this system of ODEs:

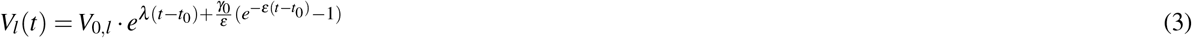

with initial conditions *V*_0,*l*_, and *γ*_0_ at time *t*_0_.

To model radiotherapy effects, at each treatment fraction delivery (*t*_*RT*_), a proportion of (1 − *S*) of the viable tumor is transferred to a dying compartment *V*_*d*_. The surviving fraction *S* is here used as a model parameter in itself, rather than as a function of radiation dose and patient specific radiation sensitivity, as described by the linear-quadratic model^21^.

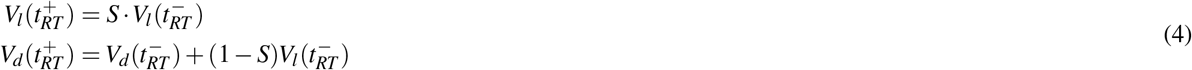

Here, 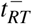 denotes time immediately before radiotherapy delivery, 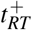 the time immediately after treatment delivery. By restricting ourselves to the same fraction size as the HFSRT treatment, the presented model provides a worst case estimate of no explicit consideration of radiation-induced immune stimulation.

We model radiation induced cell death as mitotic catastrophe^22^. Since mitotic catastrophe is a proliferation dependent process, we describe the volume change as an exponential reduction of *V*_*d*_(*t*) at rate *λ* identical to the growth rate.

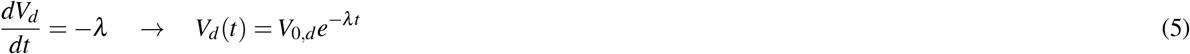

Hence, the total, observed tumour volume *V* (*t*) comprises a proliferating (*V*_*l*_(*t*)) and dying (*V*_*d*_(*t*)) population.

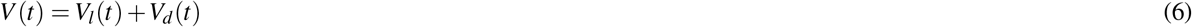

In agreement with previous results^19^ we fixed *γ*_0_ = 0.5*λ/*0.05124 and used a fixed growth rate for all patients. This results in a model comprising two patient-specific parameters (*S,ε*) as well as two patient group fixed ones (*λ, γ*_0_). A summary of the model parameters is given in Table 1.

**Table 1.** Overview of the model parameters, relevant fit bounds and range of data used for fitting (patient specific, or all patients as a whole).

| Parameter | Unit | meaning | Fit bounds | Patient specific |
| --- | --- | --- | --- | --- |
| $\lambda$ | $d^{-1}$ | Tumor growth rate in the absence of treatment | [0.017, 0.14] | - |
| $\gamma_0$ | $d^{-1}$ | Initial sensitivity to Bevacizumab and Pembrolizumab | fixed | - |
| $\varepsilon$ | $d^{-1}$ | Evolution of resistance rate | [0, 0.1] | ✓ |
| $S$ | | Radiation surviving fraction | [0, 1] | ✓ |

### 2.3 Parameter fitting and uncertainty estimation

All calculations and modelling were performed in MATLAB version 2020a. Agreement between clinically measured and simulated data was assessed by root mean squared error (RMSE). Since more than one pre-treatment volume measurements were only available for a small subset of patients (7/16), a grid search was performed to identify the most suitable growth rate for the patient population as a whole. Growth rates ranging from five to 40 days (*λ* = [0.017, 0.14]) were used for model fitting and the sum of mean, median and maximum taken over all RMSEs obtained over the full data range (including pre-treatment data) was compared. All further results are based on this optimal growth rate.

The two patient specific parameters *ε* and *S* were fitted in a sequential fashion to the relevant data: First, *ε* was fitted to the proportion of data not subject to influences of radiotherapy, which we assume to correspond to data more than four times the doubling time after HFSRT for which the dying tumor volume is less than 1*/*2^4^ of its initial value. We used MATLAB’s *fit* fsunction with default hyperparameter choices.

In a second step, under the constraint of fixed *ε*, the surviving fraction *S* was fitted to all data using a particle swarm optimizer (swarm size 100, up to 100 iterations) minimizing the sum of relative squared differences between simulated and measured tumor volume.

Parameter variation due to contouring uncertainty was estimated by bootstrapping. Each data point was shifted by multiplication with a random number drawn from a normal distribution of mean one and standard deviation 0.2 (hence assuming up to 20% uncertainty) before repeating the fit of *ε* and *S*. For data points below 2 *cm*^3^ a 0.5 *cm*^3^ uncertainty was used, independent of the recorded tumor volume to account for a minimal contouring uncertainty. Any potential negative data points were assigned to zero volume. Fifty bootstraps were calculated, and envelopes of the obtained results are shown as uncertainty bands. For all model parameters data is given as results of the undisturbed data with standard deviations over the bootstrap results. Correlation between model parameters, and between a patient’s gEUD and estimated surviving fraction *S* were evaluated with MATLAB’s *corrcoef* function.

### 2.4 Comparison of alternative treatment schedules

We first investigate the noninferiority of intermittent RT vs. HFSRT: Based on all estimated parameters the volumetric growth trajectory for each patient is simulated for five treatment fractions of the same gEUD (corresponding to the same surviving fraction per fraction) as the HFSRT delivered every six weeks. To account for treatment resistant tumors that may outgrow their pretreatment size within the six-week inter-fraction window, we also model iRT plus a single 3x gEUD (delivered in three consecutive days) boost at time of progressions. “Progression” was here defined as volume at the six-weekly assessment points exceeding the minimum measured tumor volume by more than 20%. Differences in the obtained fit parameters between patients where iRT (without boost) was inferior to HFSRT and those where it was equal or superior are compared by unpaired t-test using Matlab’s *ttest2* function.

Based on the assumption of a superior repair capacity of healthy relative to tumor tissue^23^, the intermittent treatment delivery holds the potential for reduced normal tissue toxicity at the same number of treatment fractions. In a second step, we hence investigate the potential gain in time to progression by increasing the number of intermittently delivered treatment fractions. We perform simulations for up to 13 treatment fractions.

The efficacy of any treatment schedule is evaluated based on time to progression by a Kaplan-Meier-analysis. We score the time to reach the last recorded tumour volume (cut-off volume), assuming that this provided an estimate of the patient’s maximum tolerated tumor burden. If the initial tumour volume exceeded the final recorded volume, time to reach 1.2x the initial volume was scored, since the patient left the trial at this point and no further imaging was performed. Kaplan-Meir plots were generated using the MatSurv package^24^ and log-rank p-values were calculated between HFSRT and iRT+boost treatments.

## 3 Results

### 3.1 Model fit to data

The optimal growth rate was estimated by grid search to be *λ* = 0.07 day^−1^, corresponding to a doubling time of 10 days (Figure 2A). For the data fits of all 16 patients with a fixed growth rate of 0.07 *d*^−1^ we obtained a median (minimum, maximum) RMSE of 1.8 (0.2,12.1). If considering the overall correlation of fitted and measured tumor volumina (Figure 2B) there was good agreement in terms of coefficients of determination (*R*^2^ = 0.74) which we calculated here over the logarithms of the data points to prevent an over-representation of large values in this analysis. Model fits to individual patient data are shown in the appendix (Figure S1).

**Figure 2.**
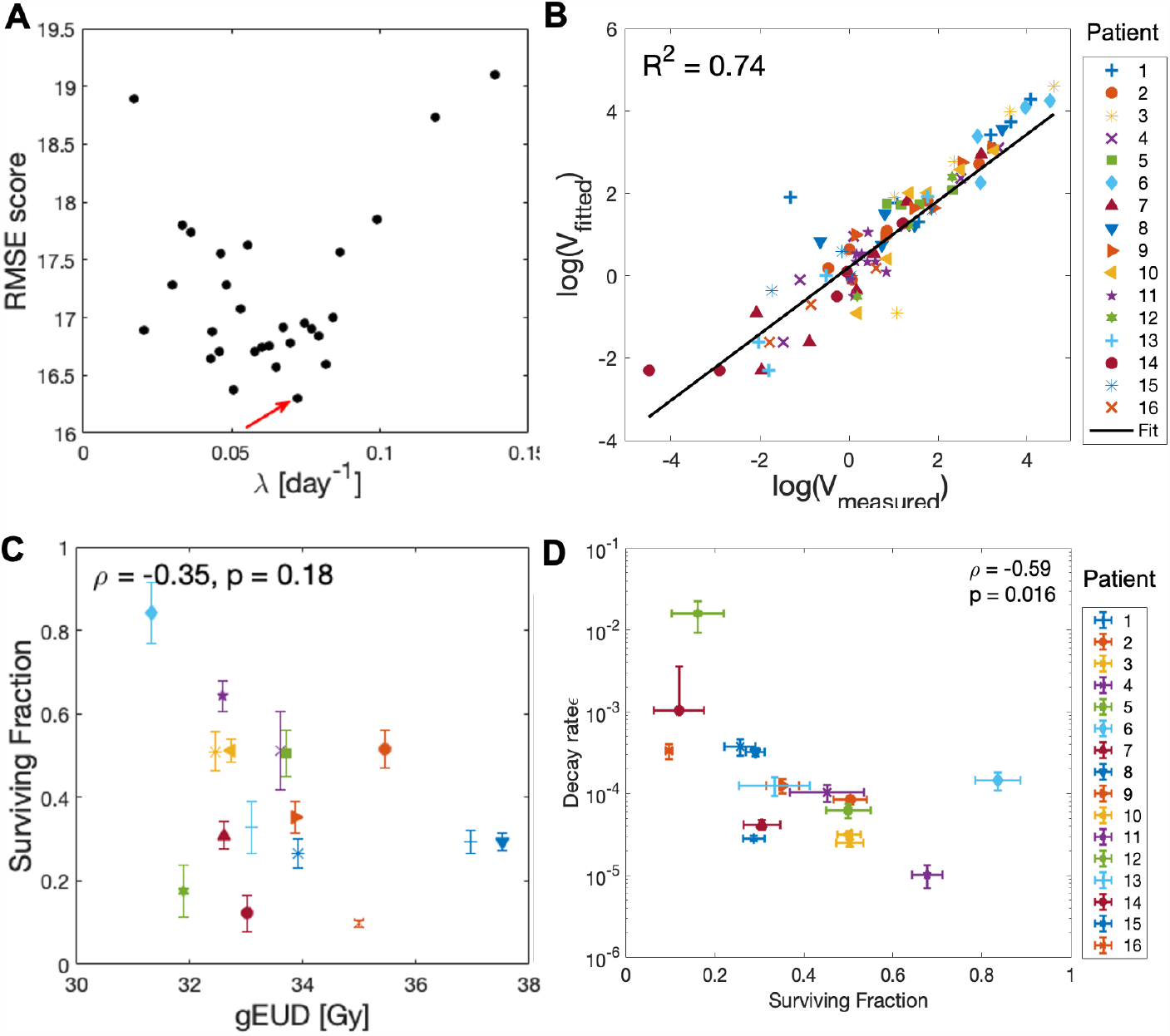
Model fit results. A) Grid search results to identify the optimal growth rate *λ* for the patient population (indicated by red arrow). Results of the sum over the median, mean and maximum RMSE are shown (denoted as RMSE score). B) Overview of the measured vs. simulated tumor volumina. C) Correlation analysis of the surviving fraction and the PTV gEUD. The Pearson correlation coefficient *ρ* and corresponding p-value *p* are given. See D for legend. D) Correlation analysis of the logarithm of the decay rate (*log*(*ε*)) and the surviving fraction. Abbreviations: RMSE: Root mean squared error, PTV: Planning target volume, gEUD: generalized equivalent uniform dose.

The obtained patient specific fit parameters, reported as median and full range, spanned a fairly large interval reflecting the biological heterogeneity and prescription dose variations: *S* = 0.34 (0.10,0.84), *ε* = 1.2 (0.1,152)·10^−4^. Interestingly, there was no correlation (*p >* 0.05) between estimated surviving fraction and planned dose in terms of gEUD (Figure 2C) or D98% (not shown) in the PTV, possibly reflecting the radiosensitivity heterogeneity of the tumors. Figure 2D shows that there was a significant negative correlation (p = 0.02) between *S* and log(*ε*) indicating that more radioresistant tumors, which are characterised by a high surviving fraction *S*, had a smaller rate of resistance development than non-RT treatments *ε*.

### 3.2 Demonstrating non-inferiority of iRT+Boost

We observed no significant difference between HFSRT and iRT (p = 0.83), or HFSRT and iRT+boost (p = 0.83) for five treatment fractions (Figure 3A). Given the small number of patients, it is, however, also important to evaluate the individually observed differences at the patient level. In 11/16 patients, iRT was equal to HFSRT, whereas time to volume cut-off was smaller for five patients. These patients (#6,8,12,14,15) were characterised by a significantly faster decay of the non-RT treatment effects (*ε*_*iRT<HFSRT*_ = 3.9 (1.5,152)·10^−4^, *ε*_*iRT>HFSRT*_ = 0.6 (0.1, 3.5)·10^−4^, p-value between log(*ε*) = 0.002)(see Figure 3B) leading to fast regrowth. There was no significant difference in the obtained radiosensitivity (indicated by *S*) of the two groups (*S*_*iRT<HFSRT*_ = 0.27 (0.12,0.84), *S*_*iRT>HFSRT*_ = 0.51 (0.10,0.64), p = 0.5) (Figure 3C). Hence, these patients would reach the cut-off volume within six weeks from the last iRT fraction.

**Figure 3.**
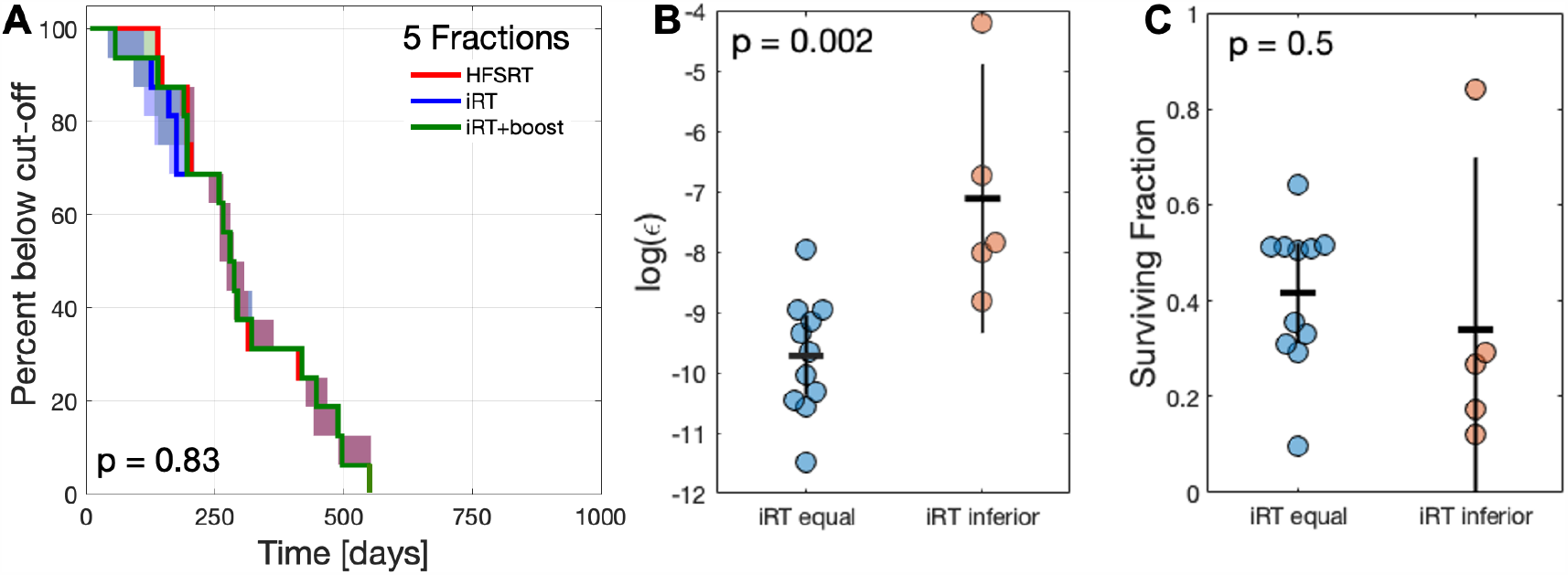
Evaluation of noninferiority of iRT+/-boost vs HFSRT. A) Kaplan-Meier plot for five treatment fractions delivered as HFSRT (red), iRT (blue) or iRT+boost (green). Shaded areas correspond to the envelope of the bootstrap estimated modelling uncertainty. The logrank test p-values is given. B) Decay rate parameter *ε* for iRT responders and non-responders. C) Surviving fraction for iRT responders and non-responders. In C) and D) the horizontal lines indicate the mean of the scores and t-test p-values are reported.

For four of these five patient time to progression could be prolonged by delivering a three-fraction boost once regrowth occurred. As such, Kaplan-Meier analysis (Figure 3A) shows that for 15 out of the 16 cases, iRT+boost was noninferior to the trial’s HFSRT treatment schedule. The single patient for whom iRT+boost was inferior to HFSRT treatment displayed the largest resistance evolution rate *ε*_*pat*19_ = 152 ± 61 · 10^−4^ day^−1^.

### 3.3 Modelling of extended intermittent treatments

Normal and tumour tissue radiosensitivity may vary strongly between individual patients. The opportunity to either continue or interrupt treatment at each of the 6-weekly evaluation time-points holds great potential for treatment personalisation. Here, the total number of delivered fractions can be adjusted leading to personalised dose escalation given the absence of acute normal tissue toxicity. In the intermittent setting, normal tissue may be capable to compensate for radiation-induced damage more effectively than the tumor, which motivates an escalation of the total delivered dose in the iRT setting. We hence simulated alternative treatments allowing up to 13 treatment fractions. As the number of fractions increases, the time to reach the cut-off volume is continuously prolonged provided the subject’s tumor shows no regrowth, which in turn increases separation of the Kaplan-Meier curves as shown for up to seven (Figure 4A), nine (Figure 4B), or eleven (Figure 4C) treatment fractions. However, given the small cohort size, differences between HFSF and iRT+boost for up to eleven intermittent treatment fractions were not significant as assessed by logrank testing (*p >*= 0.05). Only a subset of five patients (#1, 3, 7, 10, 11) would benefit further from *>* 11 treatment fractions (Figure 4D) leading to a further separation of the Kaplan-Meier plots for observation time above 550 days. However, this late separation did not change the events scored for logrank testing and hence did not further improve the p-value between HFSRT and iRT+boost treatments.

**Figure 4.**
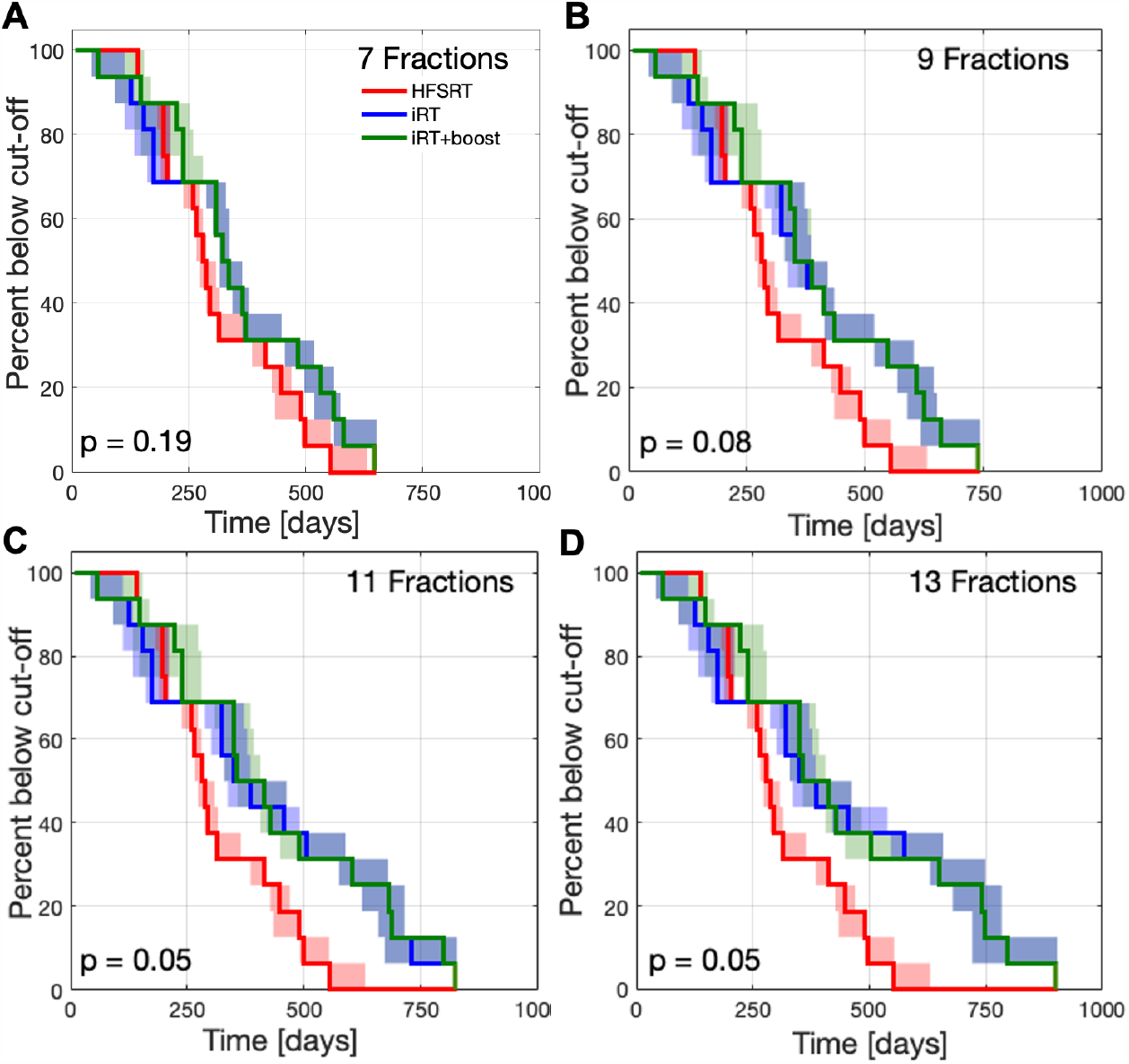
Kaplan-Meier plots for treatments with increasing maximum number of iRT fractions. Shown are fitted HFSRT (red), and simulated iRT (blue) and iRT+boost (green) results. Shaded areas correspond to the envelope of the bootstrap estimated modelling uncertainty. The logrank test p-values is given. A) Up to seven fractions. B) Up to nine fractions. C) Up to eleven fractions.

We identified four subgroups of patients with respect to their response to the different treatments: 1) HFSRT is best (# 12), 2) iRT is inferior to HFSRT, but iRT+Boost compensates this difference (# 6, 8, 14, 15), 3) iRT+boost further prolongs time to cut-off volume (# 2, 4, 5, 9, 13, 16), 4) iRT is best (# 1, 3, 7, 10, 11). Figure 5A-D shows individual growth trajectories for representative examples for each of these groups. The full set of growth trajectories is given in the supplementary material (Figure S1). The relevant evaluation of the model parameters for these sub-groups is shown in Figure 5E and F. While there was no significant difference between the radiotherapy surviving fraction, the groups differed in their fit results for parameter *ε* with small decay rates corresponding to a benefit from iRT treatment.

**Figure 5.**
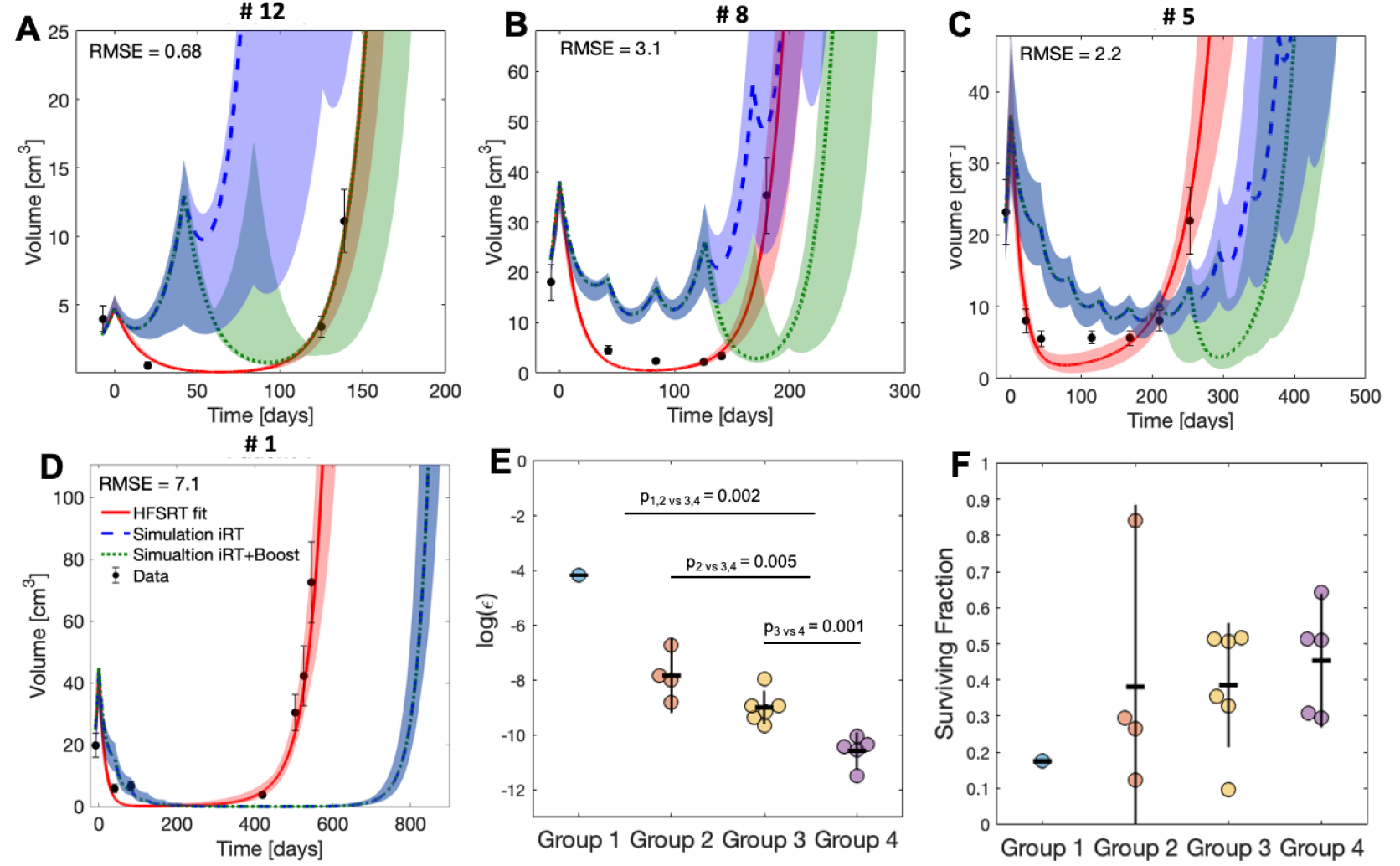
Grouping of patient response. A-D Estimated growth trajectories of representative patients for fitted HFSRT (red), and simulated iRT (blue) and iRT+boost (green) treatments with up to 11 treatment fractions. Shaded areas correspond to the envelope of the bootstrap estimated modelling uncertainty. A) Group 1, B) Group 2, C) Group 3, D) Group 4. E) Analysis of the decay rate *ε* for the different groups. t-test p-values of the logarithm of *ε* between groups are given. F) Analysis of the radiotherapy surviving fraction for the different groups. There were no significant differences.

## 4 Discussion

Based on the suggested model, it was demonstrated that an intermittent treatment plus boost should be equal to or superior than the five daily fraction HFSRT protocol for all but one of the evaluated patients. We found that the parameter characterising the rate of developing resistance to Bevacizumab and Pembrolizumab treatment was an indicator of the efficacy of iRT. Patients whose tumor developed resistance slowly, would benefit most from iRT compared to HFSRT. Based on our data it was not possible to identify these patients from pre-treatment or early evaluation time points, however it is conceivable that other biomarkers, e.g. genetic data or imaging radiomics, could be predictive of this mechanism and hence allow for stratification of patient according to the best suitable fractionation scheme. In the absence of such predictive markers, it would hence be essential to continuously monitor the tumor’s response as suggested here. We showed that iRT could possibly extend the time to tumor progression by allowing for personalization of later treatment fractions based on the observed response to the previous fraction. This could be done, as suggested here, through the option of a boost in case of progression, or by adapting the dose per fraction regimen at a personalized level as previously suggested for other tumor locations^14,20^. Fraction size variation was not addressed in this model due to the limited amount of data available. Given a more in-depth knowledge of the specific radiosensitivity parameters per tumor, for example based on genetic information, further analyses could integrate a tumor-specific radiosensitivity index^25,26^. We intended to restrict this analysis to a purely data-driven estimation of response with few patient specific fit parameters. Genetic or molecular data was not available and hence could not be accounted for.

The proposed mathematical model, despite using only two patient specific parameters, provided an acceptable fit to the data. By choosing a deliberately simple mathematical description, the number of fitted patient-specific parameters and modelling uncertainty was limited in our small patient cohort. The small cohort of only 16 patients limited the power of our analysis leading to no significant benefit of iRT+boost using more than five treatment fractions vs. HFSRT. Given the observed separation of the Kapalan-Meir plots for seven to 13 treatment fractions, our results are promising but require further evaluation on a larger set of patients. The clinical trial providing the data for this analysis demonstrated that it was safe to deliver 5x∼6 Gy as HFSRT with respect to normal tissue tolerance in recurrent HGG patients^8^. Addition of bevacizumab may have played a role in decreasing incidence of cerebral edema and radiation necrosis. Normal tissue toxicity following iRT + boost would need to be investigated for treatments comprising more than five fractions. Estimates on normal tissue complication could be related to previous results from trials investigating hypofractionated radiotherapy or stereotactic radiosurgery in combination with immunotherapy for the treatment of melanoma brain metastases^27,28^. Severe radiation-induced late side effects of brain tissue may be beyond the expected life span of recurrent high-grade glioma patients, however, acute radiation-induced side effects such as headache, seizures, intracranial haemorrhage, and brain edema^27^ should be considered. Besides these manageable toxicities, radiation necrosis may be a dose limiting factor for iRT treatments with incidence times in the order of months to few years following RT^28,29^. Acute radiation-induced toxicity may strongly correlate with the irradiated volume and dosing which together with potential normal tissue recovery between fractions makes estimations difficult. A clear advantage of the intermittent treatment approach is the option to halt further irradiation if severe acute radiation-induced toxicity occurs, which is in line with a personalized treatment approach.

Another advantage of iRT is the possibility to adapt the PTV according to the observed growth. This includes local PTV adaptations, and potential inclusion of progression sites appearing outside the primary tumor location. This type of treatment paradigm would increase treatment cost due to repeated imaging and treatment planning. Recent advances in automated treatment planning^30,31^ and the delivery of the treatment under MRI guidance with an MR-Linac^32–34^ could pose a potential solution to mitigate this limitation of intermittent treatments. Response monitoring and treatment planning steps could be combined in this scenario^35^.

It is important to clearly state the underlying assumptions made in our model which were required to limit the model’s complexity and base the simulation only on the data available: i) use of an exponential tumor growth model, ii) tumor heterogeneity was ignored, iii) bevacizumab and pembrolizumab treatments were modelled as additive effects to RT only. By maintaining rather than eradicating the tumor, growth is likely to be slower for larger tumours than those close to eradication as presented in the NCT02313272 trial. As such, modelling exponential tumor growth for both, the HFSRT and iRT scenarios would in the worst case overestimate the growth of larger tumors present following iRT. It should also be stressed that the time between fractionations for iRT (six weeks) would allow for regrowth of both, resistant and sensitive populations. Pre-clinical data and evolutionary convention^36,37^ suggest that resistant cells may display a fitness disadvantage relative to sensitive clones in the absence of the selective pressure, allowing for the sensitive subpopulation to preferentially repopulate the tumor. Hence, it would be expected that intermittent treatments, again, would provide an advantage over daily HFSRT. Additionally, intermittent treatment may hold potential for synergistic action with immunotherapy due to repeated antigen re-sampling as suggested by recent (pre-) clinical studies^38,39^. Therefore, iRT is particularly promising in combination with immune-checkpoint inhibition therapy, which suggests that the volume estimates made by our model may overestimate a real treatment scenario. In summary, all assumptions in this analysis lead to a worst case estimate of the simulated treatment response following iRT, which strengthens the results presented in this hypothesis-generating study.

Patients with recurrent HGG currently have no curative treatment options^40,41^, and radical treatment attempting tumor control may be suboptimal considering its purely palliative and life-prolonging intent. The intermittent treatment approach embraces the aim of tumor control and volume management rather than tumor eradication. This comes at the cost of potentially not levitating the tumor burden, which may affect the quality of life of the patients. For this reason, iRT should be restricted to those patients with asymptomatic recurrence or those who are neurologically stable. Our hypothesis generating modelling study provides a numerical estimate of the potential gain in time to progression for a large subgroup of patients. As such, we have demonstrated the mathematical feasibility of iRT treatments for recurrent HGG patients. This approach should be carried forward to be evaluated in a prospective clinical trial.

## 5 Conclusions

There is a critical unmet clinical need to improve response rates and overall outcome for patients with recurrent HGG. Based on a deliberately simple, worst-case estimate mathematical model we propose that intermittent radiotherapy treatments with an optional three fraction boost may be a safe (in terms of tumor control) and potentially life prolonging treatment option for this group of patients. This novel radiation treatment schedule has additional potential for personalized treatment decisions in terms of geometric dose delivery and fraction size optimization based on the observed tumor response to previous fractions.

## Data Availability

The data sets used and/or analysed in the study are available from the corresponding author on reasonable request.

## Acknowledgments

The work of this project was initiated at the 2019 Integrated Mathematical Oncology workshop at Moffitt Cancer Center for which the participation of SCB was kindly sponsored by the Moffitt Physical Sciences Oncology Center and the Integrated Mathematical Oncology Department. The authors would like to thank Dr. Renee Brady–Nicholls for providing the original code of the particle swarm optimizer.

## Author contributions statement

HE, JP and SCB devised the study and led the writing and revising of the report. SCB implemented the mathematical model and performed simulations. JP contoured MRI images and provided patient treatment information. HY provided patient MR imaging data. CJW contributed to the design of the study and mathematical model. SS was the principal investigator of NCT02313272 clinical trial. HY and SS provided clinical expertise and valuable discussion of the results. All authors reviewed and edited the manuscript.

## Additional information

### Ethical statement

All methods were carried out in accordance with relevant guidelines and regulations. The NCT02313272 (05/12/2014) was carried out at the Moffitt Cancer Center, FL between August 2015 and March 2018. All patients provided written consent and the treatment protocol and any of its amendments were approved by the institutional review board of the Moffitt Cancer Center (IRB study #: Pro00014674 and # 00000971).

## Appendix

**Figure S1.**
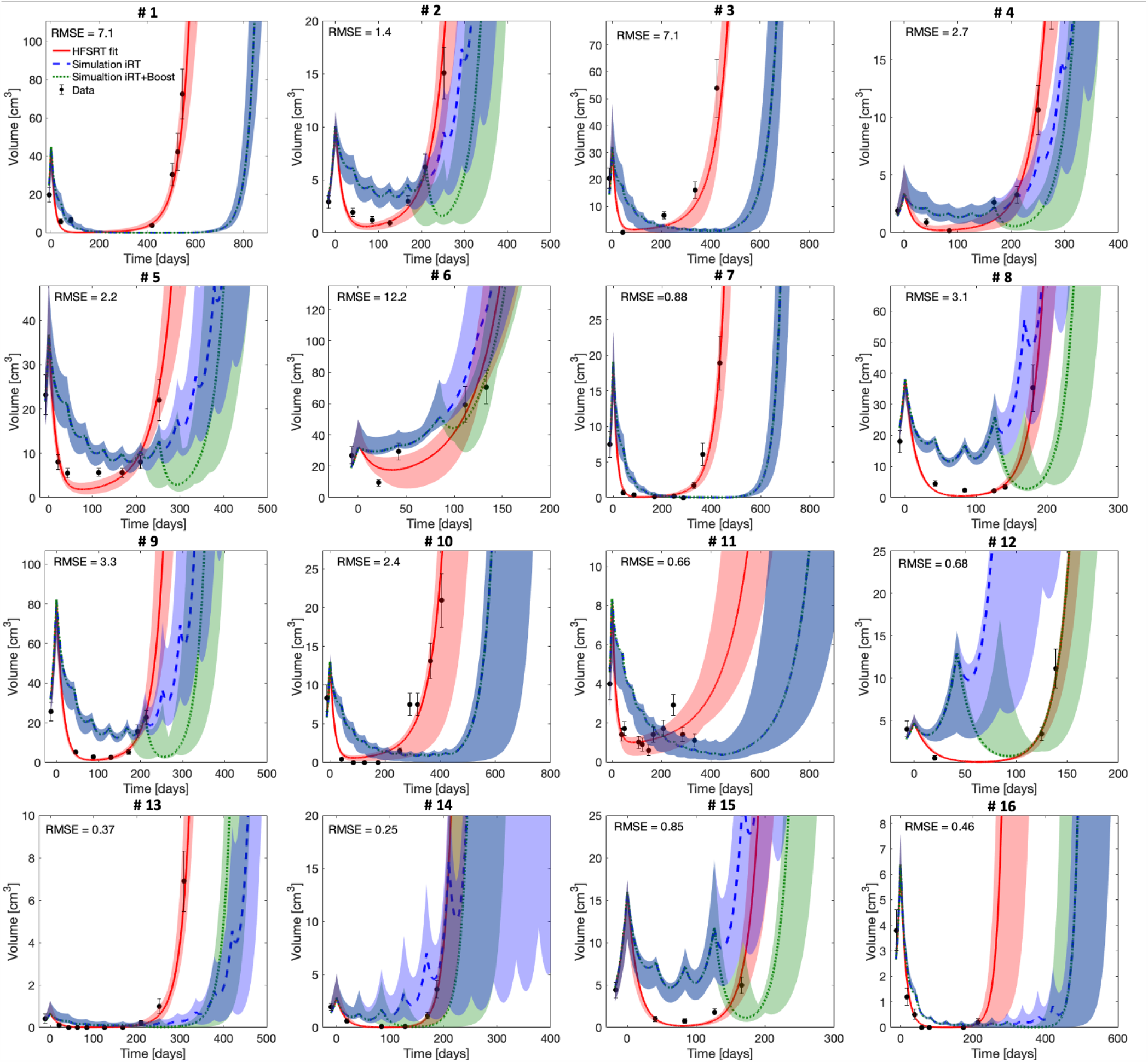
Estimated growth trajectories of all included patients for fitted HFSRT (red), and simulated iRT (blue) and iRT+boost (green) treatments with up to 11 treatment fractions. Shaded areas correspond to the envelope of the bootstrap estimated modelling uncertainty.

**Table S1.** Overview of the patient specific dosing in terms of PTV gEUD and D98%. A Lyman parameter of −10 was used for gEUD calculation. Abbreviations: PTV: Planning target volume, gEUD: generalized equivalent uniform dose, D98% near minimum dose at least received by 98% of the PTV.

| Patient | PTV gEUD [Gy] | PTV D98% [Gy] |
| --- | --- | --- |
| 1 | 37.0 | 35.9 |
| 2 | 35.5 | 34.5 |
| 3 | 32.5 | 29.3 |
| 4 | 33.6 | 30.7 |
| 5 | 33.7 | 30.6 |
| 6 | 31.3 | 29.5 |
| 7 | 32.6 | 28.8 |
| 8 | 37.5 | 34.0 |
| 9 | 33.9 | 30.5 |
| 10 | 32.7 | 31.2 |
| 11 | 32.6 | 31.0 |
| 12 | 31.9 | 30.3 |
| 13 | 33.1 | 31.1 |
| 14 | 33.0 | 30.1 |
| 15 | 33.9 | 31.9 |
| 16 | 35.0 | 31.5 |

